# The development and psychometric evaluation of the Post Activity Symptom Scale (PASS) for Myalgic Encephalomyelitis/ Chronic Fatigue Syndrome

**DOI:** 10.64898/2026.11.06.26367424

**Authors:** Mike C Horton, Sarah F Tyson, Russell Fleming

**Author notes:** Corresponding Author: Prof Sarah Tyson. Disclosure statement: None of the authors have any conflicts of interest but we note that Mr Fleming is employed by the ME Association. Funding statement: The work was funded by the ME Association, UK.

## Abstract

**Purpose:** To develop and psychometrically evaluate a new scale measuring post-exertional malaise in people with myalgic encephalomyelitis (ME)/ chronic fatigue syndrome (CFS).

**Methods:** A draft scale was co-produced with patient and clinician advisory groups. After extensive pilot testing, adults with ME/CFS completed the Post Activity Symptom Scale online and repeated it two weeks later to assess test-retest reliability and sensitivity to change. Rasch analysis assessed each domain for unidimensionality, targeting, internal reliability, item fit and local dependency. Then Rasch-transformed interval data and parametric statistics were used to assess criterion-related and discriminant validity (by comparison with measures of physical health and ME/CFS severity), test-retest reliability and minimal detectable difference.

**Results:** The item regarding ‘delay in PEM onset’ was an under-discriminating misfit anomaly which was inconsistent with the remaining PASS items. When this was removed, the PASS was a unidimensional, psychometrically robust scale. Criterion-related validity (r=0.58, p<0.001) was moderate and highly significant, discriminant validity (p<0.001) and test-retest reliability (ICC = 0.81) were excellent. The minimal detectable difference was 1.93/100. Participants found the PASS easy to complete, comprehensive and relevant.

**Conclusion:** The Post Activity Symptom Scale is a valid, reliable, sensitive measure of people of PEM in ME/CFS

## INTRODUCTION

Post-exertional malaise (PEM) is considered a cardinal symptom of Myalgic encephalomyelitis (ME)/ Chronic Fatigue Syndrome (CFS). It is a poorly understood phenomenon, described as “*the worsening of symptoms that can follow activity (of any type) that could previously be tolerated. Onset can be delayed by hours or days and has a prolonged recovery time*^1^. The mechanism of PEM is unclear but is thought to be related to a lowered aerobic threshold as part of an abnormal physiological response to, and recovery from exertion^2–4^. This means that many people with ME/CFS exceed their aerobic threshold and use anaerobic metabolism during activities in everyday life^5^. It is most clearly demonstrated during two-day cardio-pulmonary exercise testing (2-day CPET)^6,7^. On a single day CPET, people with ME/CFS demonstrate low, but relatively normal exercise capacity compared to healthy and sedentary controls. However, unlike controls, they are unable to reproduce the CPET measures when the test is repeated the following day. 2-day CPET has therefore great potential as an assessment tool. However, it is underexploited, primarily because it requires users to exercise to maximum capacity, which runs the risk of inducing PEM and precipitating serious deterioration.

Several patient-reported outcome measures (PROMs) of post exertional malaise have been published. However, none have been comprehensively psychometrically developed and tested. The best known is a collection of questionnaires by Jason and colleagues. The first, the DePaul Post-Exertional Malaise Questionnaire (DPEMQ)^8^ asks about PEM triggers, symptoms, defining features, duration, delay, and the impact of pacing over the ‘*past six months*’. Recently, Jason and Chee^9^ recommended using just two questions from this instrument for either diagnosis or outcome assessment of PEM, and that the time frame could be adapted to *“suit a variety of research or clinical contexts”*. Validation (approximating to content validity) is derived from a large survey of people with ME/CFS^10^. Criterion-related validity was established by comparison with the SF-36^9^. To the authors’ knowledge, construct validity, other aspects of validity, reliability, and sensitivity to change have not be published. The final version is available^11^ but details about how to score the questions and calculate final/total scores are not included.

The same team have also produced 5-and 10-item PEM instruments^12^ which are sub-sets of items from the DePaul Symptom Questionnaire (DSQ)^13^, which assesses all ME/CFS symptoms. The 5-item version contains descriptors of PEM from the DSQ, while the 10-item version adds questions about the nature of the PEM^12^. The authors use data from the psychometric assessment of the complete DSQ to claim good to excellent test–retest reliability based on *“Pearson’s or kappa correlation coefficients > 0.70 in the majority of items”* for the DPEMQ^14^. The only psychometric evaluation specifically of these PEM questionnaires used a German translated version of the 10-item version^15^. This reported excellent internal consistency and criterion-related validity in terms of correlation with the Patient Health Questionnaire (a measure of depression) and the Chalder Fatigue Scale (a measure of ‘tiredness’). To the authors’ knowledge, other psychometric properties have not been reported.

Davenport et al^16^ took a different approach by focussing on the impact of PEM on individuals’ activity levels and developed the PEM Activity Questionnaire. Here, the user selects three activities of daily living and rates current performance of these activities compared to a ‘good day’ and ‘before illness’ using a 0-100 scale. They also score ‘burden of functioning’ on a 0-100 scale. Reasonable test-retest reliability and responsiveness, without floor or ceiling effects have been demonstrated, but minimal detectable difference/ sensitivity to change and other aspects of validity are unknown. A considerable disadvantage is that, as it is not a standardised instrument, it could not be used as a comparative outcome in a clinical trial, as participants could select different activities to rate.

Sommerfelt et al^17^ produced the Functional Assessment of Capacity (FUNCAP) which asks about the impact of different activities on PEM. Content and face validity was demonstrated using iterative patient surveys. Construct validity is supported in terms of internal consistency/ unidimensionality and lack of floor and ceiling effects. Other aspects of construct validity, particularly redundancy/dependency have not been addressed. Known group validity between people with ME/CFS and healthy controls, and people with mild and severe ME/CFS was demonstrated. Test-retest reliability showed almost perfect replication (ICC = 0.91-0.98) but the sensitivity to change/minimal detectable difference has not been assessed. An important criticism of the FUNCAP is that items ask about the impact of completing different activities on PEM, however the response options do not consider that many people with ME/CFS carefully manage their activity (known as pacing) to prevent PEM, and there is no response option which reflects this.

Finally, a preprint publication describing the initial development and content validation of the Vienna Post-Exertional Malaise Questionnaire (VPEMQ) has been published^18^. The instrument captures four dimensions of PEM: triggers and contextual factors, symptom manifestation, functional limitations in daily life, and temporal dynamics, including onset and recovery patterns. Other psychometric properties have not yet been published,

Given that none of the available PEM PROMs assessed the nature of an individual’s PEM with comprehensively assessed psychometric properties, we developed a new one named the Post Activity Symptom Scale (PASS). Our primary aim was to co-produce a psychometrically robust tool to support clinical assessment. The work was part of larger project to develop and evaluate a clinical assessment toolkit (<u>The ME Association Clinical Assessment Toolkit (MEA-CAT) - The ME Association</u>).

## METHOD

The PASS was co-produced with people with ME/CFS and clinicians working in specialist ME/CFS services working as advisory groups who contributed to all stages of the project, as detailed in previous publications^19^. An initial draft list of items was devised from relevant literature in discussion with the advisory groups. After extensive pilot testing, a prototype scale was produced which addressed the main features of post-exertional malaise: triggers; symptoms; frequency; delay in onset; duration; and impact on day-to-day life. The questions regarding triggers and symptoms had listed different triggers/symptoms with yes/no answers and the number of triggers/symptoms was included in the total score. Users were also asked to identify the three most troublesome triggers/symptoms of PEM. This was considered important information for clinical assessment and to open discussions about managing PEM but would not contribute to an overview of PEM severity. Consequently, they were not scored and did not contribute to the total score. Similarly, users were asked whether they experienced ‘warning signs’ of PEM as this was important to understand the patient’s PEM but was not scored as it relates to the nature of PEM, rather than its severity. The other questions used a simple Likert scale answer format.

We resolved to avoid using the term ‘post-exertional malaise’ in the title and the body of the scale. This was because some people find the terms ‘exertion’ and ‘malaise’ problematic as they cannot exert themselves, and malaise was considered redolent of malingering. Furthermore, feedback from the clinicians was that patients often did not understand the terms and they were difficult to explain. So, we referred to a *‘worsening of symptoms when you overdo it’.* The timescale for questions was “over the last month, in keeping with the rest of the MEA-CAT.

The prototype PASS was converted into an online survey with alternative formats for those who could not use a computer screen. Adults in the UK diagnosed with ME/CFS were recruited via publicity in ME/CFS support groups, the 25% ME group (a charity supporting people with severe ME/CFS) and the ME Association’s social media and publicity channels.

The COSMIN study design checklist for patient-reported outcome measurement instruments was used to structure the analysis and reporting^20^. Rasch Measurement Theory (RMT) was used as the basis for scale development and psychometric testing of the internal construct validity of the PASS^21^. This was followed by classic theory testing of other aspects of validity, test-retest reliability and sensitivity to change/minimal detectable differences. The Rasch Model is a unidimensional measurement model that provides a template against which measurement instruments can be tested, due to its satisfaction of fundamental measurement assumptions^22,23^. When items (questions) are summed together into a single total score that represents an underlying trait, RMT provides a unified framework for assessing several aspects of internal construct validity. The intention is to assess how each individual item operates and contributes towards the total score, and to highlight any measurement anomalies within an item set. Rasch analysis of the data was completed with RUMM2030 software^24^, using the partial credit model^25^. The Rasch analytic process included several standard tests of fit, covering both the scale and item-levels.

- All items were assessed individually for fit to the Rasch model within the item set, confirming that each item contributes satisfactorily to the underlying construct. Misfit indication guides were taken as significant Bonferroni-adjusted chi-square p-values or when standardised (z-score) fit-residuals fall outside ±2.5, alongside visual inspection of the item-characteristic curves^26,27^. An emphasis was placed on under-discriminating items (high positive fit residuals) which imply a lack of contribution to the overall score, and that these items may measure something other than the construct in question^28^.
- Unidimensionality was evaluated by a series of t-tests^29^ with multidimensionality indicated if independent subsets of items delivered significantly different person estimates, with lower bound 95% CI percentage of significantly different t-tests >5%.
- Response category functioning determined whether the items’ response structure was operating as intended, where functional response category structures are indicated by sequential response thresholds (the crossover points between subsequent response categories) on the underlying logit scale^30^.
- Tests of local dependency (LD) were used to assess whether any items were more closely related than is explained by the underlying construct, which can potentially indicate a structural overlap or redundancy within the item set. An initial LD indicator was taken as any residual correlation (Q3 value) greater than 0.2 above the average residual correlation, although the significance of this cut point is variable depending on various attributes of the dataset^31^.
- Item bias was assessed through uniform and non-uniform differential item functioning (DIF) testing by sex and age group, with significant DIF indicated at a Bonferroni adjusted ANOVA p-value.
- Scale targeting was assessed graphically through the relative distribution of item and person locations. Reliability indices were taken as the person separation index (PSI) and Cronbach’s alpha values^26^.
- It is known that large sample sizes tend to overpower the fit statistics within RUMM^32^. Therefore, the full sample was randomly split into two and run as a cross-validation sample to ensure replication of results and to reduce the risk of capitalising on a chance finding. The cross-validation subsamples were used to assess indications of individual item fit and DIF, whereas the complete sample was used to assess indications of targeting, reliability (PSI and Cronbach’s alpha), LD, unidimensionality, response category functioning, and floor and ceiling effects.

Prior to inspection of the complete PASS item set, the triggers and symptoms items were assessed separately. These items list different triggers/symptoms with yes/no responses, and the total number of triggers/symptoms is included in the total PASS score. Both the triggers and symptoms items essentially act as super-items, with several separate component items contributing to the individual triggers and symptoms item scores. Therefore, a separate subscale analysis was carried out for the ‘triggers’ subset (11 dichotomous items) and the ‘symptoms’ subset (11 dichotomous items) to ensure that each of the component items are validly contributing to the total triggers and symptoms scores that are utilised in the PASS.

Criterion-related and discriminant validity, test-retest reliability and minimal detectable difference (MDD) were evaluated using a classic theory testing approach. Rasch-converted data were used, where the ordinal PASS raw scores were transformed into interval-level (0-100 scales) for easy interpretation^33^. This provides the benefits of continuous data properties and allows parametric analyses to be performed. The conversion chart used for this transformation is available in Supplementary File 1 and is freely available for others to use.

As well as the PASS, participants completed the physical health (physical function and role limitations) section of the SF36 (SF36-PH)^34^ and NICE ME Severity scale^1^ to assess criterion-related and discriminant validity. Pearson Correlations between the PASS and the SF36-PH assessed criterion-related validity. Discriminate validity used analysis of variance (ANOVA) to establish whether PASS score differed in with ME illness severity. Both tested the hypothesis that PEM severity would be greater in people with more severe ME/CFS and disability.

Two weeks later, participants repeated the PASS to assess test-retest reliability and MDD, and stated whether their ME/CFS had been the same, better or worse over the previous two weeks. Data from those who replied ‘the same’ were compared with their original scores using Intra-Class Correlations (ICC, two-way random effects model), while MDD was calculated from the standard error of measurement of the intra-class correlation (ICC) where:

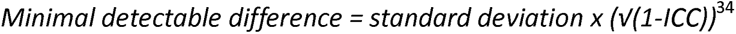

## RESULTS

The participants’ characteristics are detailed in Table 1. They were predominantly middle-aged women with moderately ME/CFS of long duration.

**Table 1.**
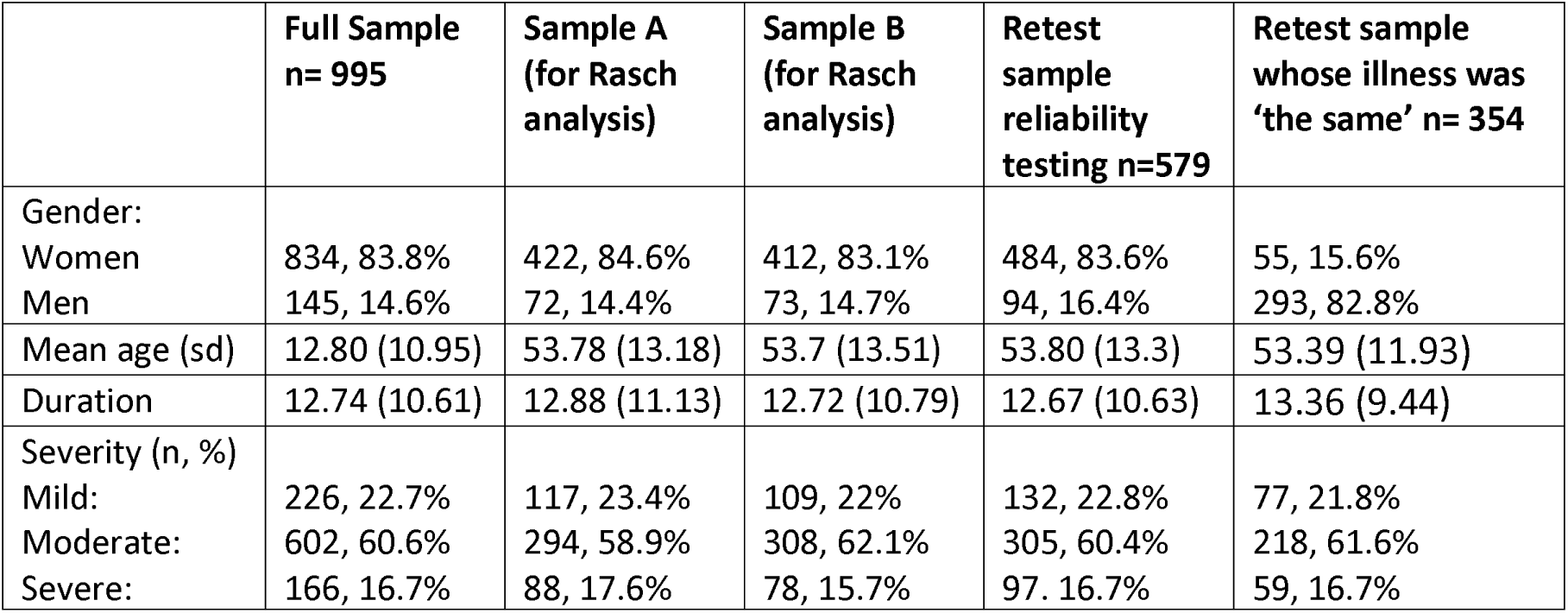
Participant characteristics of the different samples.

|  | **Full Sample n= 995** | **Sample A (for Rasch analysis)** | **Sample B (for Rasch analysis)** | **Retest sample reliability testing n=579** | **Retest sample whose illness was 'the same' n= 354** |
|---|---|---|---|---|---|
| Gender: |  |  |  |  |  |
| Women | 834, 83.8% | 422, 84.6% | 412, 83.1% | 484, 83.6% | 55, 15.6% |
| Men | 145, 14.6% | 72, 14.4% | 73, 14.7% | 94, 16.4% | 293, 82.8% |
| Mean age (sd) | 12.80 (10.95) | 53.78 (13.18) | 53.7 (13.51) | 53.80 (13.3) | 53.39 (11.93) |
| Duration | 12.74 (10.61) | 12.88 (11.13) | 12.72 (10.79) | 12.67 (10.63) | 13.36 (9.44) |
| Severity (n, %) |  |  |  |  |  |
| Mild: | 226, 22.7% | 117, 23.4% | 109, 22% | 132, 22.8% | 77, 21.8% |
| Moderate: | 602, 60.6% | 294, 58.9% | 308, 62.1% | 305, 60.4% | 218, 61.6% |
| Severe: | 166, 16.7% | 88, 17.6% | 78, 15.7% | 97. 16.7% | 59, 16.7% |

### Rasch Analysis

#### Triggers and Symptoms

All the dichotomous items in the ‘triggers’ and ‘symptoms’ questions contributed sufficiently towards the total scores, with no standout misfit anomalies (see Supplementary Files 2 and 3). In the trigger subset, ‘pain’ did display a misfit value outside of the normally accepted range (Fit Residual = −3.09) in one of the sub-samples, but this was not consistent across both cross-validation samples. Both item sets displayed evidence of unidimensionality (series of t-tests = 0.74% significant for ‘triggers’ and 0.47% for ‘symptoms’), although it should be noted that the power of the series of t-tests is limited. No significant LD was noted for triggers. For symptoms, potentially significant LD was noted between ‘fatigue’ and ‘brain fog’, but no further action was taken as all items are combined into a single super-item for the PASS, thus accounting for any potential LD within the item set. Across all samples, the ‘physical’ trigger and the ‘fatigue’ symptom were most commonly reported, and the ‘hormonal’ trigger and the ‘allergy’ symptom were least common.

#### Other items of the PASS

The initial PASS individual item fit and LD are reported in Table 2. It was observed that the ‘Delay’ item was a standout misfit anomaly across both cross-validation sub-samples, displaying consistently significant underdiscrimination. This is illustrated in the item characteristic curve (ICC, Figure 1) where the observed data (black dots) form a much flatter curve than is expected by the model (solid grey line). It can also be seen that the magnitude of the misfit (Chi^2^ value) is much greater for ‘Delay’ item than the other items, again indicating its presence as an anomaly within the item set. The ‘Triggers’ and ‘Symptoms’ items also displayed a degree of misfit, with fit statistics outside of the commonly accepted range. However, these items display a slight over-discrimination, meaning that they still contribute to the underlying construct, and the magnitude of the misfit is not as great.

**Figure 1.**
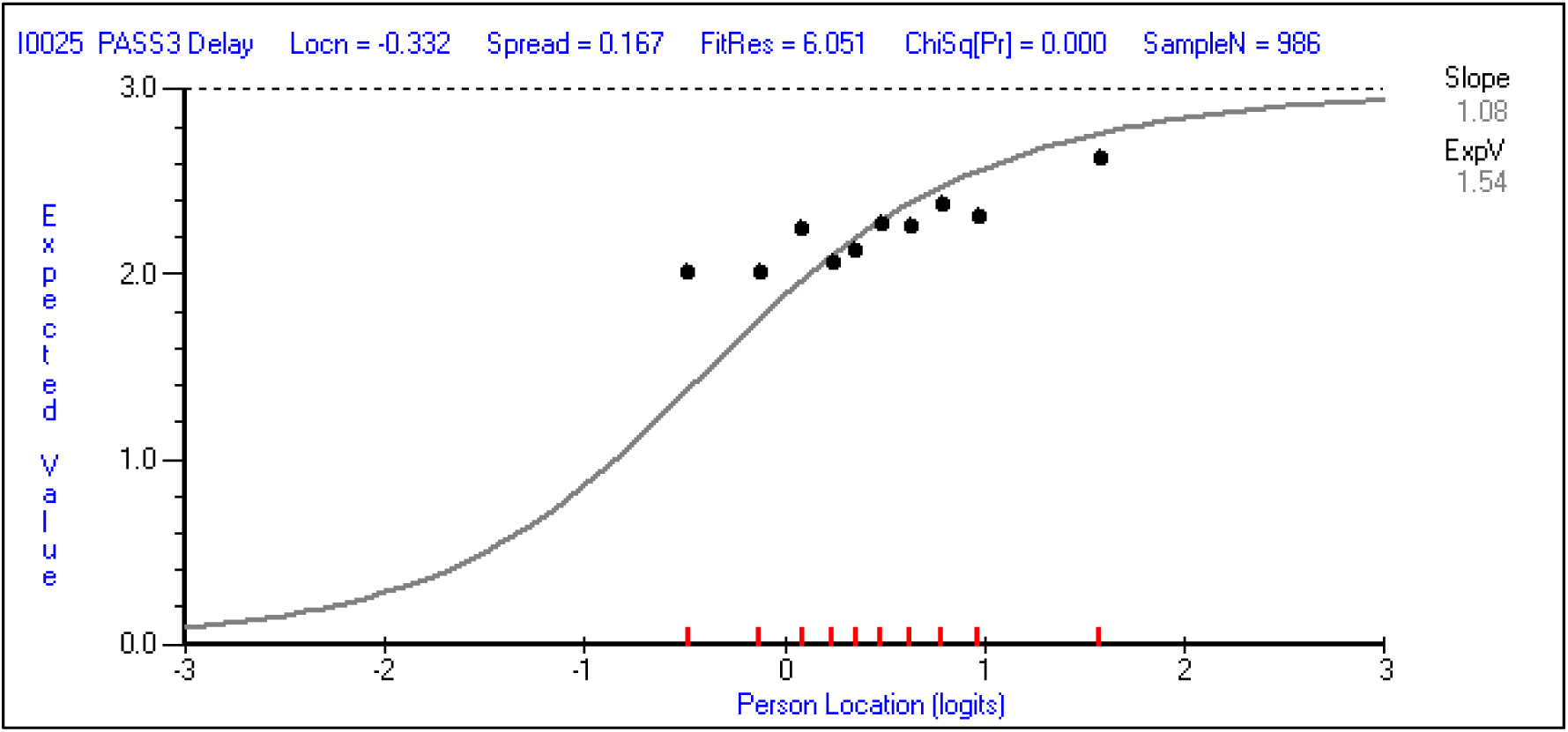
Item Characteristic Curve of the under-discriminating ‘Delay’ item.

**Table 2.**
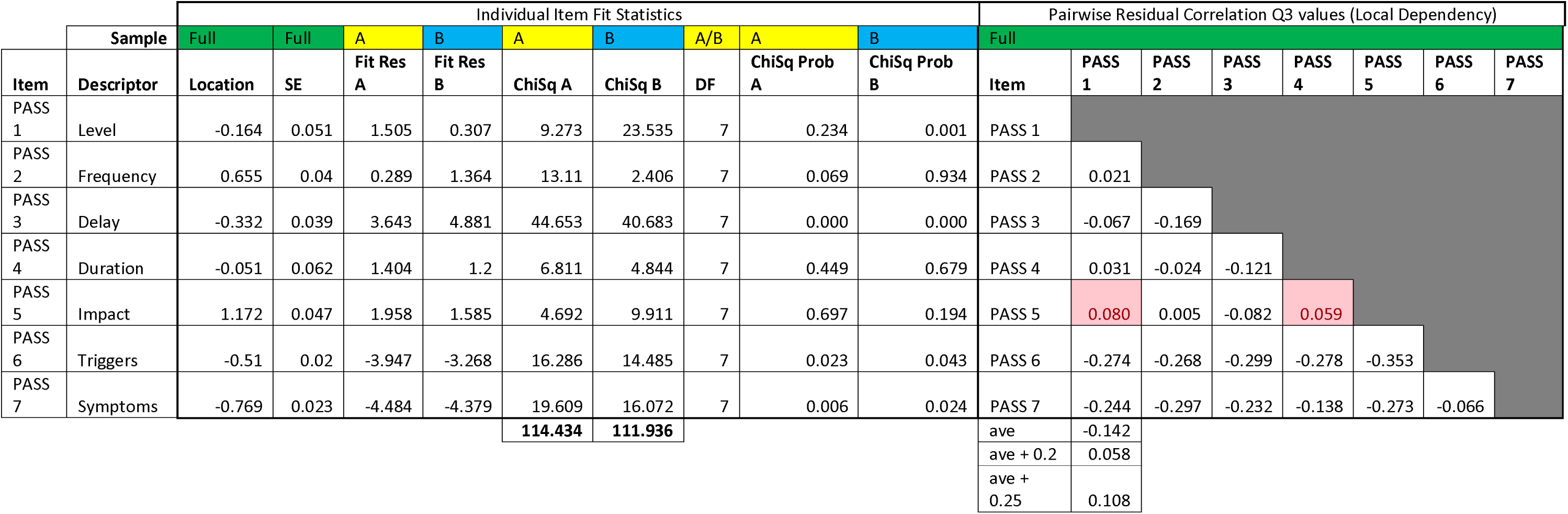
Initial individual item fit and local dependency of the PASS item set.

Despite the clear ‘Delay’ anomaly, the item set as whole appeared to be unidimensional (series of t-tests = 3.75% significant). Response category thresholds were ordered for all items except for the ‘Delay’ item, although the extent of the disordering was borderline. There was some potential LD observed between two of the 21 pairwise residual correlations. This was largest between the ‘Level’ and ‘Impact’ items. There was no apparent DIF-by-Gender or DIF-by-Age. Each of the cross-validation samples indicated one significant DIF, but on different items. Targeting of the item set showed a slight right skew of 0.58 logits, indicating that, on average, the sample had a slightly higher level of PEM burden than is represented by the items (Figure 2). The reliability indices were moderate (person separation index = 0.65, Cronbach’s Alpha = 0.6) but there were no floor or ceiling effects.

**Figure 2.**
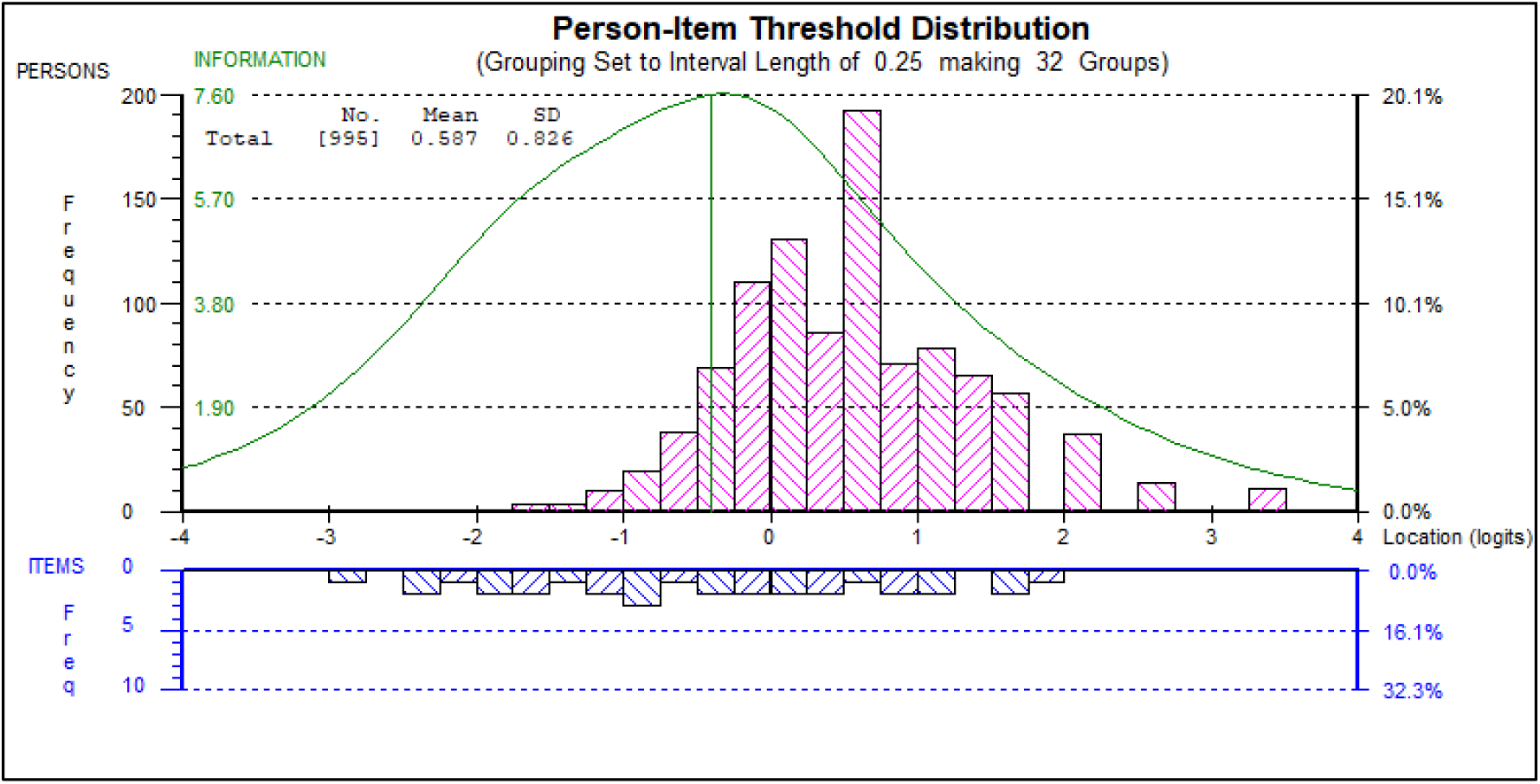
Targeting plot of the final PASS item set. This plot shows the relative distributions of the ME/CFS sample (above the x axis) and the measurement points of the PASS (below the x axis), when placed on the common logit scale. A slight right skew of 0.59 logits can be seen, indicating that, on average, the sample has a slightly higher level of PEM burden than is represented by the items. There are no floor or ceiling effects.

Due to the ‘Delay’ item representing a clear anomaly, it was removed from the item set. Note that this doesn’t necessarily mean that it is a bad item, per se, it just means that it does not work alongside the other items of the PASS. Also note that various rescoring options were explored prior to removal, but it continued to remain anomous. As the delay in onset of PEM is an important clinical issue, particularly when it comes to activity management, it was retained within the PASS but was not scored as part of the item set.

Following removal of the ‘Delay’ item, there were no further clear standout anomalies in the PASS individual item fit or LD within the item (Table 3). None of the items consistently misfitted in terms of the chi-square fit, although ‘Triggers’, ‘Symptoms’ and ‘Impact’ still displayed a degree of Fit Residual misfit, with fit statistics slightly outside of the commonly accepted range. However, the magnitude was borderline.

**Table 3.** Individual item fit and local dependency of the final PASS item set (with ‘Delay’ item removed)

|  |  | Individual Item Fit Statistics |  |  |  |  |  |  |  |  | Pairwise Residual Correlation Q3 values (Local Dependency) |  |  |  |  |  |  |
|---|---|---|---|---|---|---|---|---|---|---|---|---|---|---|---|---|---|
|  | **Sample** | Full | Full | A | B | A | B | A/B | A | B | Full |  |  |  |  |  |  |
| **Item** | **Descriptor** | **Location** | **SE** | **Fit Res A** | **Fit Res B** | **ChiSq A** | **ChiSq B** | **DF** | **ChiSq Prob A** | **ChiSq Prob B** | **Item** | **PASS 1** | **PASS 2** | **PASS 4** | **PASS 5** | **PASS 6** | **PASS 7** |
| PASS 1 | Level | -0.182 | 0.052 | 2.844 | 0.995 | 10.826 | 20.663 | 7 | 0.146 | 0.004 | PASS 1 |  |  |  |  |  |  |
| PASS 2 | Frequency | 0.668 | 0.041 | 0.564 | 2.068 | 10.944 | 11.753 | 7 | 0.141 | 0.109 | PASS 2 | 0.005 |  |  |  |  |  |
| PASS 4 | Duration | -0.058 | 0.063 | 1.599 | 1.796 | 15.735 | 8.887 | 7 | 0.028 | 0.261 | PASS 4 | 0.020 | -0.048 |  |  |  |  |
| PASS 5 | Impact | 1.195 | 0.048 | 2.643 | 2.663 | 4.171 | 2.198 | 7 | 0.760 | 0.948 | PASS 5 | 0.073 | -0.014 | 0.048 |  |  |  |
| PASS 6 | Triggers | -0.653 | 0.022 | -3.383 | -2.643 | 16.822 | 8.132 | 7 | 0.019 | 0.321 | PASS 6 | -0.311 | -0.333 | -0.325 | -0.394 |  |  |
| PASS 7 | Symptoms | -0.97 | 0.024 | -3.574 | -3.609 | 13.267 | 17.726 | 7 | 0.066 | 0.013 | PASS 7 | -0.274 | -0.348 | -0.166 | -0.302 | -0.159 |  |
|  |  |  |  |  |  | **71.765** | **69.359** |  |  |  | ave | -0.169 |  |  |  |  |  |
|  |  |  |  |  |  |  |  |  |  |  | ave + 0.2 | 0.031 |  |  |  |  |  |
|  |  |  |  |  |  |  |  |  |  |  | ave + 0.25 | 0.081 |  |  |  |  |  |

The item set as whole was unidimensional (series of t-tests = 3.46% significant) and response category thresholds were ordered for all items. There was borderline LD between two of the 15 pairwise residual correlations, primarily ‘Level’ and ‘Impact’. No apparent DIF-by-Gender or DIF-by-Age were reported; the cross-validation samples indicating some significant DIF, but on different items. Item set targeting showed a slight right skew. The reliability indices remained moderate, although both the person separation index of 0.68 and the Cronbach’s Alpha of 0.63 improved. Floor or ceiling effects were absent.

The results of the Rasch analysis and other psychometric properties of the PASS are summarised in Table 4 including the results for the full sample and cross-validation samples for both for the original and revised item set (i.e. with and without the ‘delay’ item).

**Table 4.**
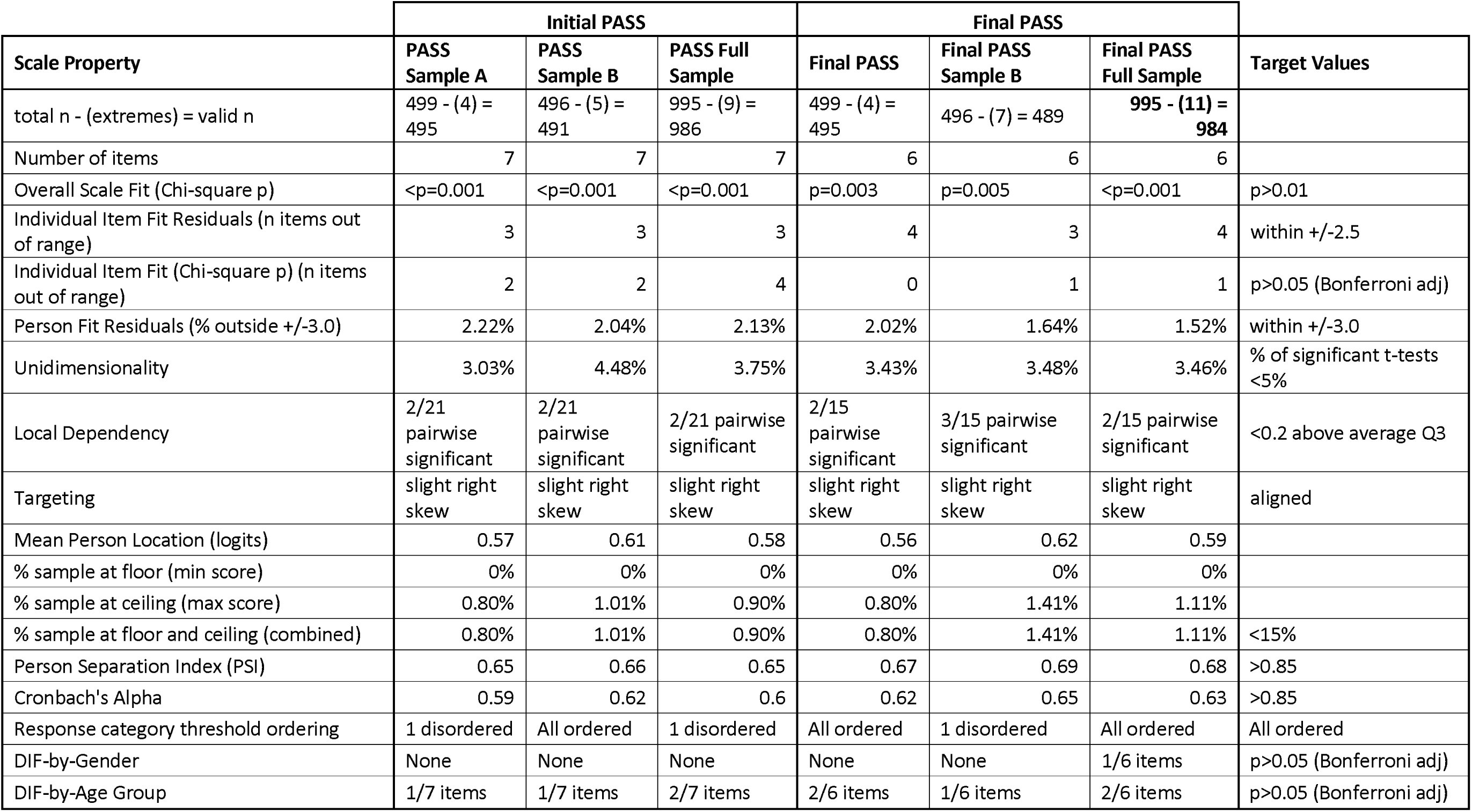
PASS Psychometric properties summary table.

#### Criterion-related and discriminant validity

Criterion related validity between SF36-PH and the PASS was moderate but highly significant (r=0.580, p<0.001). Discriminant validity was strong in that there were highly significant differences (p<0.001) in PASS score between participants who rated their ME/CFS as mild, moderate or severe. The PASS demonstrated excellent test-retest reliability (ICC = 0.814, p<0.001) with a minimal detectable difference of 1.93.

## DISCUSSION

The results show that the Post Activity Symptom Scale (PASS) is a unidimensional, psychometrically robust, reliable, sensitive assessment of post exertional malaise in people with ME/CFS. The final version is found in Supplementary file 4 or can be downloaded <u>here</u>. It is freely available for use.

Although the fit of the final PASS item set to the Rasch model is not perfect, there were no clear anomalies. We therefore believe it is justifiable to sum the PASS item set into a total score that is sufficient for the purpose it is being used for. None of the items under-discriminate in a way that suggests they are measuring another separate trait, therefore validating the unidimensional total score of the PASS scale. Furthermore, all analyses were repeated across cross-validation samples, thus reducing the risk of acting on chance findings. Additionally, the PASS contains two items that consist of counts of separate triggers and symptoms. These types of counts are usually assumed to validly represent the construct through the total count, but this is rarely tested. Within this study, these items are treated as ‘super-items’ within the context of the Rasch analysis, but each has been investigated separately to ensure all the separate individually listed triggers and symptoms legitimately contribute to the total score of the corresponding super-item.

As an ordinal scale, the PASS’s main value may be to describe and classify people’s experience of PEM. However, the reliability and minimal detectable difference are strong so it could also be used as an outcome measure, or to monitor change over time when Rasch transformed interval data are used. This approach is novel and offers greater measurement precision compared to the use of ordinal data. This is due to Rasch-transformed scores’ greater sensitivity, particularly when the distribution of scores is not located in the mid-range of the scale but include the endpoints of the scale range. The greater sensitivity to change increases statistical power so substantially smaller sample sizes would be needed to detect a change. A further advantage of using interval level data is that it enables PASS data to be included in parametric analyses with other continuous data (e.g. physiological or biomedical markers) which would increase the breadth and depth of such investigations. There have been few previous publications comparing raw ordinal and Rasch transformed data. Most recently, similar effects were reported in our work to develop and evaluate The Index of ME Symptoms (TIMES) which is another element of the MEA-CAT^36^.

The main strength of this study lies in its large sample size, co-production approach, rigorous use of Rasch methods, and utilisation of Rasch-transformed interval data for the classic theory testing. The main limitation lies in the representativeness of the sample. We specifically limited recruitment to adults and so the PASS will need to be validated for children and young people with ME/CFS before implementation in this population. However, the demographics broadly reflect other adult ME/CFS studies with convenience samples^37–39^. Thus, we are confident that the views expressed here are reasonably representative of the ME/ CFS community. However, although we recruited people with a self-reported ‘formal’ (I.e. from a health care professional) diagnosis of ME/CFS, we did not confirm the diagnosis via health records. This is because the recording of ME/CFS in both primary and secondary care is notoriously inaccurate, incomplete and inconsistent^40–41^. However, it does mean that we may have recruited some people who would prove to have other diagnoses if thoroughly assessed or accurately recorded.

## Data Availability

All data produced in the present study are available upon reasonable request to the authors

